# Longitudinal analysis of UK Biobank participants suggests age and APOE-dependent alterations of energy metabolism in development of dementia

**DOI:** 10.1101/2022.02.25.22271530

**Authors:** Jun Liu, Najaf Amin, William Sproviero, Matthias Arnold, Richa Batra, Bruno Bonnechere, Yu-Jie Chiou, Marco Fernandes, Jan Krumsiek, Danielle Newby, Kwangsik Nho, Jun Pyo Kim, Andrew J Saykin, Liu Shi, Laura Winchester, Yang Yang, Alejo J Nevado-Holgado, Gabi Kastenmüller, Rima F Kaddurah-Daouk, Cornelia M van Duijn

## Abstract

Experimental models shows that bioenergetic homeostasis changes with increasing age based on apolipoprotein E (*APOE*) gene. However, such link with dementia remains unclear in population. We used H^1^-NMR metabolome in blood from 118,021 random-selected participants in UK Biobank (n=118,021 individuals), and identified 56 metabolites associated with the risk of dementia. In the participants without developing dementia during follow-up, 82% (46/56) metabolites are also associated with reaction time, and dementia shares metabolite signatures with total brain volume. We found that incident Alzheimer’s disease (AD) is associated with energy metabolism-related metabolites, i.e, β-hydroxybutyrate, acetone, and valine, whose concentrations in blood are influenced by age and *APOE* genotype. Valine shows a declined trajectories after a plateau at age around 60 years which is in parallel with body mass index. Moreover, we found associations of AD with valine and β-hydroxybutyrate in brain tissue are different as their associations in the periphery, which implies the key role of transports in regulating the energy metabolism of AD. Our study provides strong evidence in population level that the onset of AD in *APOE4* carriers is regulated by the impaired energy balance in the brain.

## Background

Dementia is developing into one of the major worldwide population health problems^1^. The most common diseases underlying dementia are Alzheimer’s disease (AD) and vascular dementia (VaD)^1^. Obesity, diabetes, and hypertension have been identified as cardiometabolic risk factors for dementia, both AD and VaD^1^. However, the association of weight and blood pressure with future dementia are age-dependent. While midlife obesity and hypertension predict incident dementia, the levels of these risk factors decrease when approaching the onset of disease, most likely as a consequence of the insidious disease process^1–7^. As a result, by the time of diagnosis, patients often present with lower weight or blood pressure^2–7^. The loss of weight and decrease in blood pressure has been shown to begin up to 10 years before the diagnosis of dementia^8^.

The mechanism that explains loss of weight remains to be elucidated. From a clinical and epidemiological perspective, a key question is why glucose levels in the blood remain high in patients even they develop dementia. Glucose levels are strongly correlated to weight and are expected to parallel weight loss^8,9^. In the past decade, we and other groups have uncovered metabolite signatures in blood during the pathogenesis of dementia or AD^9–12^. These involve a wide variety of metabolites, including branched-chained amino acids (BCAAs), glucose, triglycerides, fatty acids, carnitines, and lipid particles^9–12^. Of note, the relation of BCAAs with dementia and AD is different from its metabolic effect observed in diabetes: while BCAA levels in blood are increased in patients with diabetes^13^, low levels of valine are associated with the risk of dementia and AD^10,14–16^. From a molecular perspective, a gap in our knowledge is how the peripheral metabolites change over time.

*Apolipoprotein E*4 (APOE4)* variant is the most important genetic factor of AD, the major type of dementia. A recent study of aged transgenic *APOE* mice on energy metabolism may shed light on the observed non-linear association of weight and dementia across life course. They found that the transgenic *APOE4* mice showed bioenergetic deficits that involve the tricarboxylic acid (TCA) cycle at old age^17^. The TCA cycle is required by the brain which cannot utilize lipids^18^. Under normal conditions, the brain relies on catabolism of glucose for producing adenosine triphosphate (ATP). Although patients with AD have high levels of glucose in the blood, cerebral metabolic rates for glucose are decreased, in particular in carriers of the *APOE4* variant^19^. Besides glucose, ketone bodies are another key energy resource of brain. They can be produced by astrocytes in the brain for the neurons as the most important alternative fuel when the brain needs more energy^20^. Alternatively, the liver can produce ketone bodies and these can be transported to the brain through the blood circulation (liver-brain axis)^20^. Besides ketone bodies, BCAAs can also be utilized by brain cells as energy substrates in the TCA cycle and as essential nitrogen donors essential for brain. Though BCAAs metabolism is also in the astrocytes, unlike ketone bodies, they cannot be produced by body and can be obtained from food intake only. While there is increasing evidence for a collapse of energy metabolism in the brain of dementia patients^21^, there is no direct evidence in large-scale population studies for crosstalk between the periphery and brain for dementia or AD.

Here, we aim to explore the metabolic signature of dementia using a prospective study in 118,021 random-selected participants of the UK Biobank with 249 metabolic measures assessed by nuclear magnetic resonance (H^1^-NMR) (referred to as metabolites; Supplementary Table 1)^22^. We identify multiple metabolites associated with dementia or AD and show that energy metabolism in the periphery is changed in patients developing AD. We find that those who are developing AD and those with early pathology signs at magnetic resonance imaging (MRI) or cognitive testing have increased blood levels of ketone bodies and other metabolites associated with energy metabolism. Our study further finds an interplay between age, *APOE* and metabolites involved in energy metabolism over the adulthood that may be relevant for our understanding of the pathogenesis, prevention, and treatment of dementia, especially of AD.

## Results

### Incident dementia associates with 56 metabolite measures

Table 1 and Supplementary Table 2-3 summarize the characteristics of the 118,021 participants included in the current study. During follow-up until September 2020 (mean follow-up years = 10.2), 1,364 participants developed dementia: 553 were diagnosed with AD, 298 with VaD, and 465 with unspecified dementia. Since the vast majority of dementia cases were aged 60 years or over (n=1,188, 87.1%), the analyses for dementia and its subtypes were restricted to this age group (Supplementary Table 3-4). We found 151 out of the 249 (61%) metabolites significantly associated with dementia (false discovery rate, FDR<0.05), when we adjusted for basic confounders, including ethnicity, age, sex, body mass index (BMI), fasting time, assessment centre, as well as batch and spectrometer during the NMR measurements (null model; Supplementary Table 5). The number of significant metabolites reduced from 151 to 56 (FDR<0.05), when further adjusting for smoking status, alcohol intake frequency, education and drug intake, which were previously found to be associated with the majority of the Nightingale metabolites (discovery model; Supplementary Table 5)^23^. The metabolites include ketone bodies (β-hydroxybutyrate and acetone), acetate, glucose, citrate, BCAAs (valine, leucine and total BCAA), fatty acids (omega 3, omega 6, Polyunsaturated fatty acids (PUFA) and Linoleic acid (LA)), six very large high-density lipoprotein (HDL) particles, M_HDL_TG, seven small HDL particles, 19 very-low-density lipoprotein (VLDL) particles, total triglycerides and ten percentages of VLDL, low-density lipoprotein (LDL) or HDL particles (Figure 1). *APOE* genotype is the major determinant of the metabolites assessed in the platform with 222 metabolites associated with *APOE4* and 233 metabolites associated with *APOE2* (FDR<0.05; Supplementary Table 6). To adjust for pleiotropic metabolic effects of *APOE* that are not related to dementia, we adjusted for *APOE* as a sensitivity analysis, reducing the number of significant metabolites (FDR<0.05) to 43 with six HDL particles and seven percentages excluded (Supplementary Table 5). The findings were similar when analyzing Europeans only or including participants with all ethnicities (correlation coefficient = 0.99 across all models; Supplementary Figure 1; Supplementary Table 7). When looking into the major subtypes of dementia, VaD is not significantly associated with any metabolites (FDR<0.05) but is nominally significantly associated with small HDL and very large HDL particles (S_HDL_P/L/PL/C/CE and XL_HDL_P/L/C/CE/FC; p<0.05) which were not found to be associated with AD (Figure 1; Supplementary Table 8); while AD is significantly associated with β-hydroxybutyrate, and acetone (FDR<0.05) and borderline significantly with valine (p=0.001, FDR adjusted p = 0.08).

**Table 1.** Description of the overall study population involved in the current study

| Variables | Total (n = 118,021) | Non-event (n = 116,657) | Patients diagnosed with dementia during follow-up (n = 1,364) |
| --- | --- | --- | --- |
| Baseline age (years), Mean ± SD | 56.51 ± 8.09 | 56.42 ± 8.08 | 64.19 ± 4.96 |
| Male, n (%) | 54093 (45.83) | 53338 (45.72) | 755 (55.35) |
| BMI (kg/m<sup>2</sup>), Mean ± SD | 27.44 ± 4.78 | 27.43 ± 4.78 | 27.66 ± 4.86 |
| Genetic risk score (GRS) of AD, Mean ± SD | 0.045 ± 0.006 | 0.044 ± 0.006 | 0.048 ± 0.007 |
| GRS without APOE, Mean ± SD | 0.042 ± 0.003 | 0.042 ± 0.003 | 0.043 ± 0.003 |
| Family history of dementia, n (%) | 13755 (11.65) | 13460 (11.54) | 295 (21.63) |
| Ethnicity, n (%) |  |  |  |
| White | 111666 (94.62) | 110348 (94.59) | 1318 (96.63) |
| Asian | 2319 (1.96) | 2302 (1.97) | 17 (1.25) |
| Black | 1792 (1.52) | 1777 (1.52) | 15 (1.1) |
| Chinese | 344 (0.29) | 344 (0.29) | 0 (0) |
| Mixed | 666 (0.56) | 659 (0.56) | 7 (0.51) |
| Others | 1234 (1.05) | 1227 (1.05) | 7 (0.51) |
| APOE genotypes, n (%) |  |  |  |
| e33 | 68727 (58.62) | 68183 (58.84) | 544 (40.3) |
| e22 | 693 (0.59) | 690 (0.6) | 3 (0.22) |
| e23 | 14370 (12.26) | 14273 (12.32) | 97 (7.19) |
| e24 | 3099 (2.64) | 3062 (2.64) | 37 (2.74) |
| e34 | 27576 (23.52) | 27047 (23.34) | 529 (39.19) |
| e44 | 2768 (2.36) | 2628 (2.27) | 140 (10.37) |
| Education, n (%) |  |  |  |
| A levels AS levels or equivalent | 13382 (11.34) | 13240 (11.35) | 142 (10.41) |
| CSEs or equivalent | 6407 (5.43) | 6375 (5.46) | 32 (2.35) |
| College or University degree | 38260 (32.42) | 37967 (32.55) | 293 (21.48) |
| NVQ or HND or HNC or equivalent | 8289 (7.02) | 8174 (7.01) | 115 (8.43) |
| O levels GCSEs or equivalent | 25280 (21.42) | 25038 (21.46) | 242 (17.74) |
| Other professional qualifications | 6216 (5.27) | 6135 (5.26) | 81 (5.94) |
| None | 20187 (17.1) | 19728 (16.91) | 459 (33.65) |
| Smoking status, n (%) |  |  |  |
| never | 64503 (54.65) | 63877 (54.76) | 626 (45.89) |
| previous | 41011 (34.75) | 40431 (34.66) | 580 (42.52) |
| current | 12507 (10.6) | 12349 (10.59) | 158 (11.58) |
| Frequency of alcohol use, n (%) |  |  |  |
| Daily or almost daily | 23925 (20.27) | 23642 (20.27) | 283 (20.75) |
| Less than once a week | 36214 (30.68) | 35697 (30.6) | 517 (37.9) |
| Once or twice a week | 30454 (25.8) | 30154 (25.85) | 300 (21.99) |
| Three or four times a week | 27428 (23.24) | 27164 (23.29) | 264 (19.35) |
| Baseline diabetes, n (%) | 7278 (6.21) | 7058 (6.09) | 220 (16.28) |
| Baseline hypertension, n (%) | 60471 (54.04) | 59523 (53.82) | 948 (72.48) |
| Baseline cardiovascular disease, n (%) | 47051 (40.13) | 46187 (39.86) | 864 (63.95) |
| Baseline dyslipidemia, n (%) | 42715 (41.99) | 42217 (41.98) | 498 (42.82) |
| Cognitive functions |  |  |  |
| Fluid intelligence score, Mean ± SD (n=36,715) | 5.98 ± 2.17 | 5.99 ± 2.17 | 4.98 ± 2.02 |
| Reaction time (ms), Mean ± SD (n=114,746) | 559.07 ± 117.79 | 558.34 ± 117.15 | 622.89 ± 151.38 |
| MRI regions |  |  |  |
| Hippocampal volume (cm<sup>3</sup>), Mean ± SD (n=9,585) | 3.83 ± 0.45 | 3.84 ± 0.45 | 3.40 ± 0.50 |
| Total brain volume (cm<sup>3</sup>), Mean ± SD (n=9,585) | 1161.44 ± 110.45 | 1161.50 ± 110.50 | 1123.05 ± 78.63 |
| White matter hyperintensities (cm<sup>3</sup>), Mean ± SD (n=9,191) | 5.00 ± 6.53 | 4.98 ± 6.47 | 17.35 ± 23.26 |

**Figure 1.**
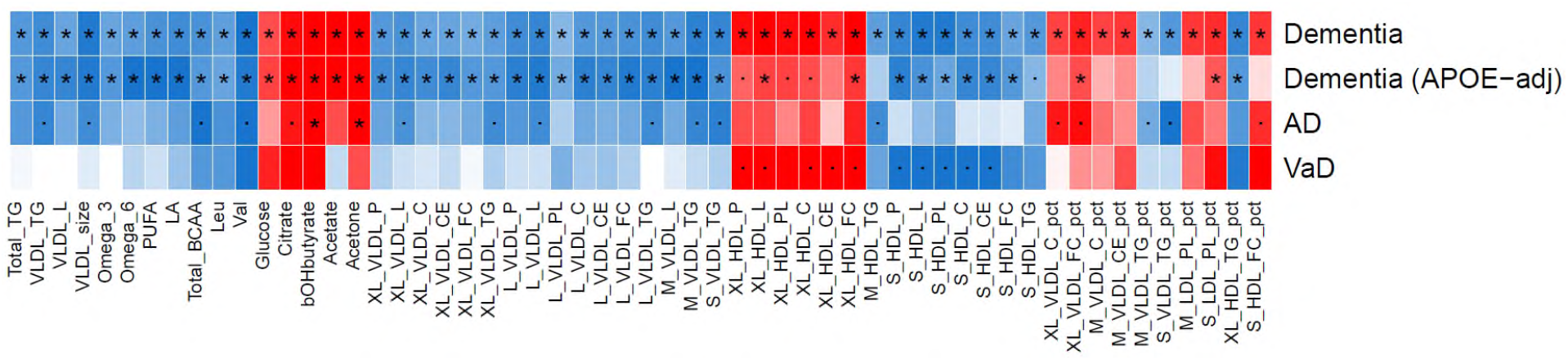
Metabolite associations of incident dementia or major subtypes. Comparison of the metabolite associations of incident dementia in different models (n = 1,188/49,843 for dementia, n = 494/50,537 for Alzheimer’s disease (AD), n = 269/50,762 for vascular dementia (VaD)). It shows only the significant metabolites (FDR-adjusted p value < 0.05, n = 56) in the discovery model with adjustment of ethnicity, age, sex, body mass index, fasting time, assessment center, and technical variables during the NMR measurement, i.e., batch and spectrometer, smoking status, alcohol intake frequency, education and drugs. **APOE-adj** is the sensitivity analysis in dementia with additionally adjusted for *APOE*. The association results of the full set of 249 metabolites are shown in Supplementary Table 5. Dot (.) denotes p < 0.05 and star (*) denotes FDR-adjusted p value < 0.05. Red: positive association; blue: negative association; the depth of the color presents the strength of effect. The codinsg of the metabolites are presented in Supplementary Table 1.

### Dementia shares metabolite signatures with its early pathology in non-demented participants

We next studied the relationship of the metabolite associations with dementia and with early pathological changes of dementia in the participants who did not develop dementia during follow-up (based on the discovery model; Figure 2; Supplementary Table 9). We studied five endophenotypes including three MRI-derived measures (hippocampal volume, total brain volume and white matter hyperintensities), fluid intelligence reflecting verbal and numerical reasoning ability, and reaction time speed to cognitive tasks. To this end, we compared the metabolite signatures of dementia and the endophenotypes in Figure 2A, 2C, 2E, 2G, 2I, which plot the effect estimate per standard error for each metabolite with the endophenotype (X-axis) and with dementia (Y-axis). As expected the effect estimates of hippocampal volume and total brain volume were negatively associated with those of dementia, while the effect estimates of reaction time were positively associated with those of dementia (Figure 2A, 2C, 2G). The metabolites from the Nightingale platform are strongly correlated within and across classes (Supplementary Figure 2). To take the correlation and the joint effects of the metabolites into account, we repeated the comparison of effects using 27 principal components (PCs), which explained over 95% of the full set of the 249 metabolites (Figure 2B, 2D, 2F, 2H, 2J). The metabolic effects captured by associations of these PCs with dementia significantly correlated with the metabolic effects of total brain volume (r= -0.47, p=0.012, Figure 2D) and reaction time (r=0.60, p=0.001, Figure 2H); correlations with the profile of hippocampal volume were borderline significant (r=-0.35, p=0.08; Figure 2B). For white matter hyperintensities (r= -0.26, p=0.19; Figure 2F) and fluid intelligence score (r=-0.04, p=0.83; Figure 2J), we did not find evidence for a joint metabolic signature (Supplementary Figure 3 for sensitivity analysis; Supplementary Table 9-10). Focusing on individual metabolites, we see a strong overlap for reaction time with dementia: 46 (82%) of the 56 metabolites associated with dementia were also significantly associated with reaction time in the non-demented participants (FDR<0.05). In detail, Figure 2G shows that the metabolites most strongly positively associated with both the risk of dementia and long reaction time included acetone, β-hydroxybutyrate, four very large HDL particles, and two percentages, i.e, S_LDL_PL_pct and XL_VLDL_PC_pct. Valine, small HDL, median VLDL and large VLDL particles are associated with both reduced risk of incident dementia and short reaction time. None of the 56 metabolites associated with dementia were associated with total brain volume at FDR significance (Figure 2C). However, several dementia-related metabolites are associated with total brain volume at nominal significance, including total BCAAs, valine, leucine, LA, omega 6, small HDL particles, citrate, glucose and very large HDL particles.

**Figure 2.**
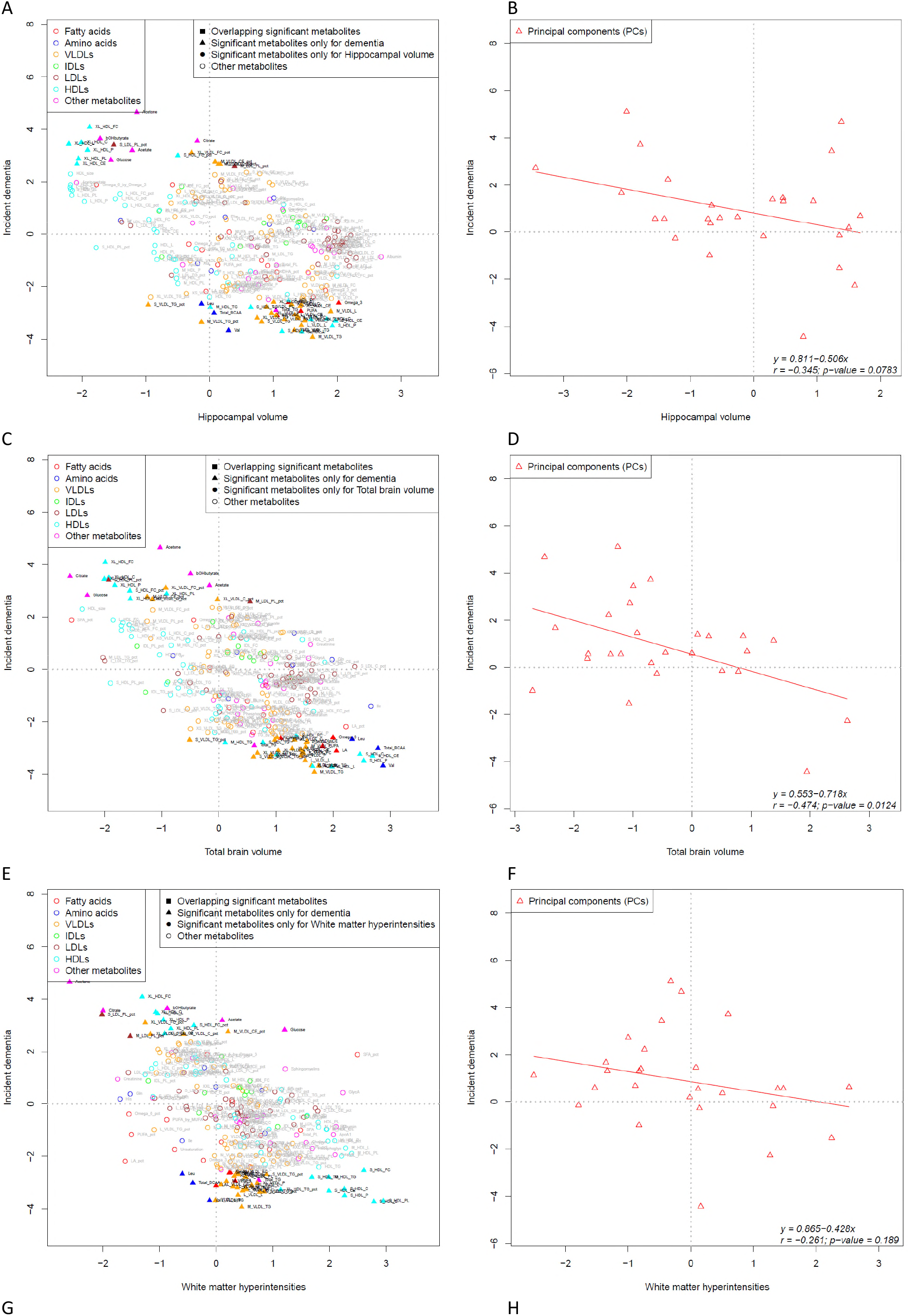
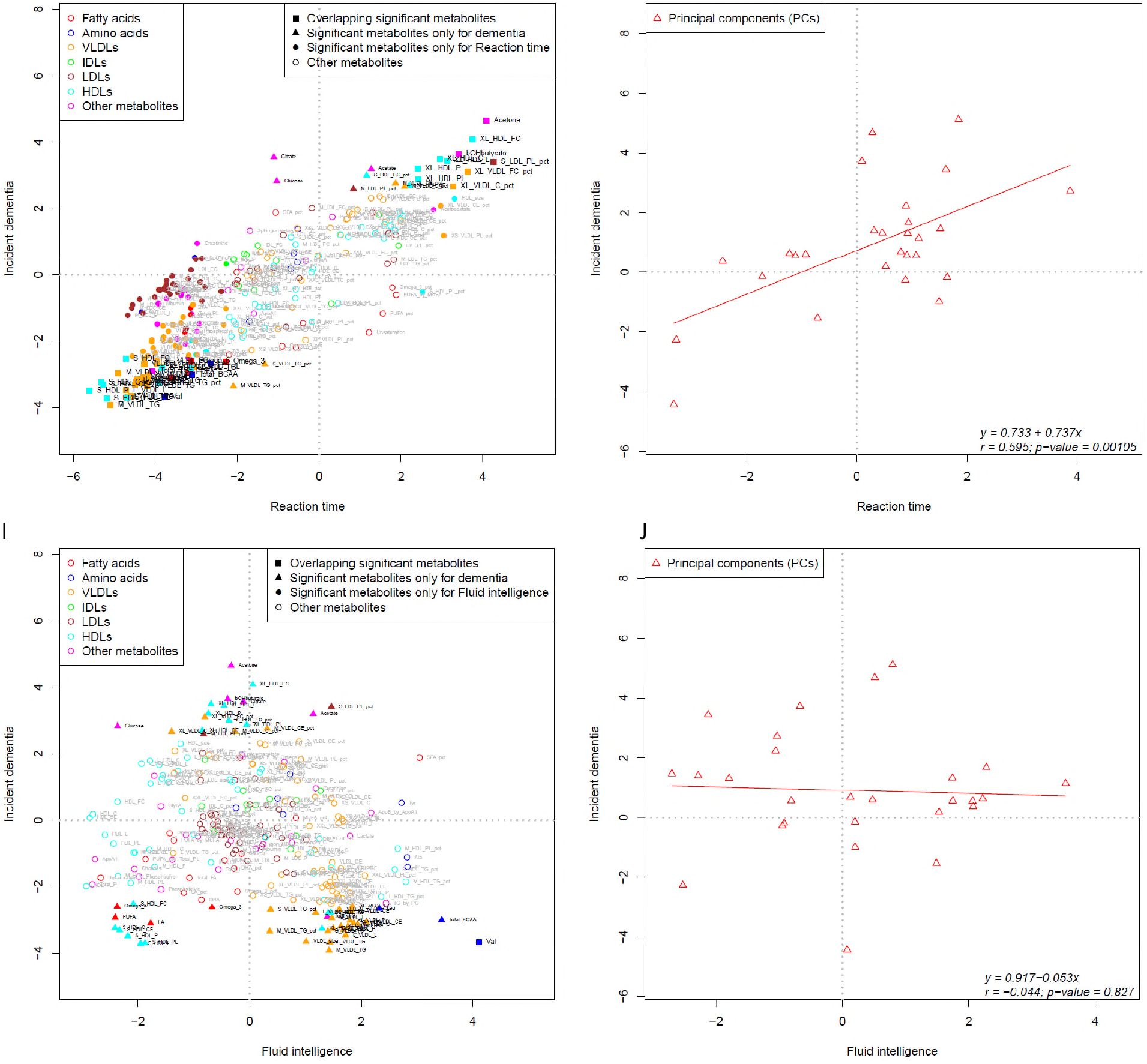
Integrating metabolite signatures of dementia and its early pathological variables, including MRI and cognitive function testing, in the participants who did not develop dementia during follow-up. The first 27 principal components (PCs) which explain over 95% of the 249 metabolites were used to overcome the high correlation among the metabolites. The effect estimates per standard error with incident dementia and the endophenoypte based on the discovery model were presented in the x-axis and y-axis respectively. Linear regression and Pearson’s correlation test were used to fit the linear model of the metabolite association pattern through PCs. bOHbutyrate: β-hydroxybutyrate; Val: valine. The coding of other metabolites are presented in Supplementary Table 1.

We further extended the integrating analysis to AD which has smaller sample size but is still powerful to identify associated metabolites (Figure 1). Although the metabolite signatures of AD and total brain volume were not significantly correlated (r=-0.27, p=0.18; Figure 3A, 3B), we still identified that valine is associated with both AD (borderline significant) and total brain volume (p_AD_ =0.001; p_total brain volume_ =0.004). Moreover, we found the metabolic signature of reaction time was significantly correlated with those of AD (r=0.45, p=0.018; Figure 3D). Figure 3C shows that both reaction time and AD were significantly positively associated with β-hydroxybutyrate (p_AD_=4.7×10^−6^; p_reaction time_=6.4×10^−4^) and acetone (p_AD_=1.7×10^−4^; p_reaction time_=4.2×10^−5^). Valine was the leading metabolite negatively associated with both AD (p=0.001) and reaction time (p=1.6×10^−4^; Figure 3C). The integrating results of AD and early pathology highlighted the importance of energy metabolism-related metabolites in AD, including β-hydroxybutyrate, acetone and valine.

**Figure 3.**
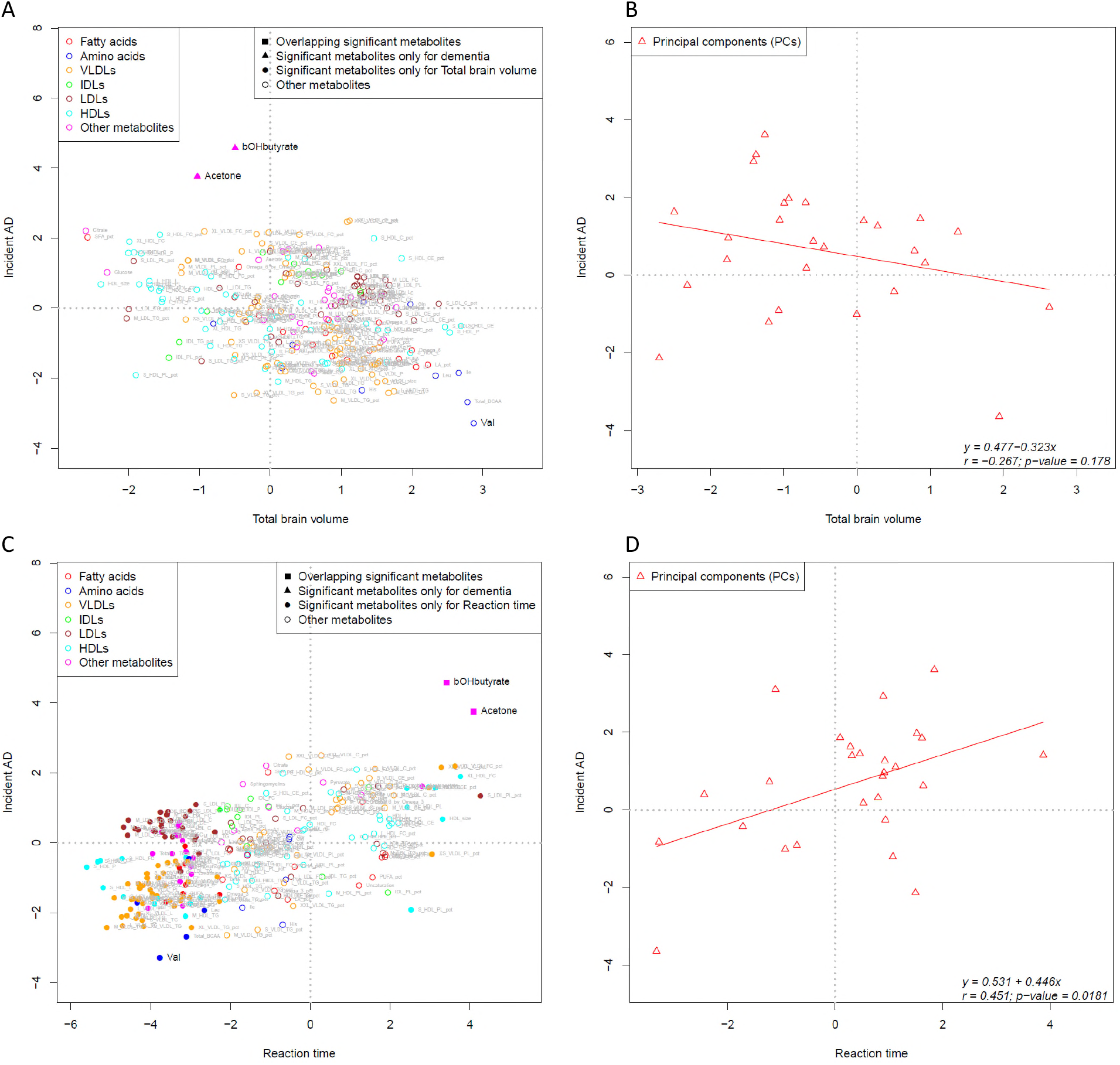
Integrating metabolite signatures of Alzheimer’s disease (AD), and total brain volume and reaction time in the participants who did not develop dementia during follow-up. The first 27 principal components (PCs) which explain over 95% of the 249 metabolites were used to overcome the high correlation among the metabolites. The effect estimates per standard error with incident dementia and the endophenoypte based on the discovery model were presented in the x-axis and y-axis respectively. Linear regression and Pearson’s correlation test were used to fit the linear model of the metabolite association pattern through PCs. bOHbutyrate: β-hydroxybutyrate; Val: valine. The coding of other metabolites are presented in Supplementary Table 1.

### Age and *APOE* involve in the association of AD and energy metabolism-related metabolites

Building upon the phenomenon of inverse association of weight and dementia across life course and the animal based studies that suggest an imbalance in energy metabolism in aged *APOE4* animals^17^, we next figured the distribution of the concentrations of these energy metabolism-related metabolites across the large age range from 37 to 73 years covered by UK Biobank and stratified by *APOE*. These analyses were performed in the population who did not develop dementia (n=116,657; Figure 4). We found that the concentrations of β-hydroxybutyrate and acetone increased with age across all the *APOE* genotypes (Figure 4A, 4B). Across all ages studied, the levels of β-hydroxybutyrate in the carriers of *APOE4* allele were significantly higher than in the *APOE33* carriers (beta=0.042, p=1.7 × 10^−9^). The levels of β-hydroxybutyrate in the *APOE2* carriers were lower compared to the levels in *APOE33* carriers (beta=-0.032, p=4.9×10^−4^). Yet, β-hydroxybutyrate levels in blood increased more strongly over age in *APOE2* carriers (beta _interaction with age_=0.003, p=0.018; Figure 4A). Acetone levels did not differ significantly depending on *APOE* genotypes (Figure 4B). For valine, we found that the levels across the age range were higher in *APOE2* carriers and increased with age (beta=0.037, p=3.4 × 10^−5^), while valine in *APOE4* carriers were lower (beta=-0.015, p=0.03) and started to decrease after age 60 years (p= 8.0 × 10^−30^; Figure 4C). Figure 4D shows that after age 57-58 years, the levels of BMI in *APOE4* carriers started to decline with age (p= 1.1 × 10^−15^) and the interaction between *APOE4* and age is significant (beta _interaction with age_ = -0.015; p=0.0002). The findings in Figure 4 remained significant when adjusting for confounding effect of other covariates (Supplementary Figure 4). As a similar pattern was shown between valine and BMI (Figure 4C, 4D), we further validated the correlation of valine and BMI in the non-demented participants, and showed that valine is strongly associated with BMI overall (beta=1.24; p=1.06 × 10^−21^), in those aged under 60 years (beta=1.94; p=4.8×10^−6^) and those aged 60 years or older at baseline (beta=1.12; p=6.76×10^−17^).

**Figure 4.**
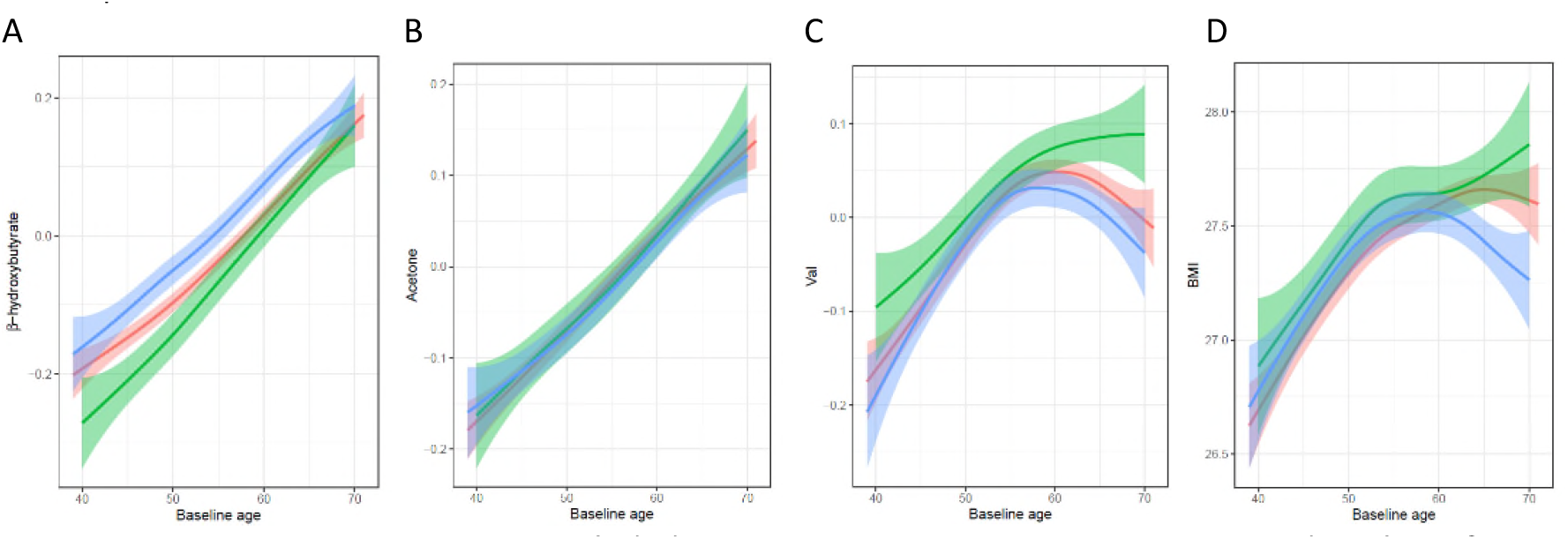
The distribution of metabolites and BMI over the life course by APOE in the participants who did not develop dementia. Participants carrying *APOE24* were excluded; participants carrying *APOE33* were used as the reference group (red colour). *APOE2* carriers are presented in green colour and *APOE4* carriers in blue colour. Generalized additive model was used to smooth the lines in the figure. Linear regression was performed to model the lines, and showed below. Age: baseline age; age^2^: the square of baseline age; * interaction. The metabolites are coded based on Supplementary Table 1. (A) β - hydroxybutyrate = - 0.016 + 0.013 × age (p = 1.5 × 10 ^-142^) - 0.032 × *APOE2* (p = 4.9 × 10 ^-4^) + 0.042 × *APOE4* (p = 1.5 × 10 ^-9^) + 8.2 × 10 ^-5^ × age^2^ (p = 0.09) + 0.003 × age\**APOE2* (p = 0.018) + 6.6 × 10 ^-5^ × age\**APOE4* (p = 0.94) (B) Acetone = - 0.004 + 0.010 × age (p = 2.0 × 10 ^-98^) + 0.003 × *APOE2* (p = 0.70) - 0.042 × *APOE4* (p = 0.55) + 8.0 × 10 ^-6^ × age^2^ (p = 0.87) + 0.0004 × age\**APOE2* (p = 0.72) - 0.0005 × age\**APOE4* (p = 0.57) (C) Valine = 0.035 + 0.0047 × age (p = 3.3 × 10 ^-22^) + 0.037 × *APOE2* (p = 3.4 × 10 ^-5^) - 0.015 × *APOE4* (p = 0.033) - 0.0005 × age^2^ (p = 8.0 × 10 ^-30^) + 0.0008 × age\**APOE2* (p = 0.49) - 0.001 × age\**APOE4* (p = 0.27) (D) BMI = 27.6 + 0.026 × age (p = 9.7 × 10 ^-30^) + 0.11 × *APOE2* (p = 0.01) - 0.043 × *APOE4* (p = 0.20) - 0.002 × age^2^ (p = 1.1 × 10 ^-15^) - 0.003 × age\**APOE2* (p = 0.58) - 0.015 × age\**APOE4* (p = 1.9 × 10 ^-4^)

β-hydroxybutyrate was the metabolite most strongly associated with AD, age, and *APOE*. A further intensive sensitivity analysis was performed in the association of AD and β-hydroxybutyrate. After considering the effect of co-morbidities, other potential biomarkers or drugs related to β-hydroxybutyrate levels (drugs were selected based on the metabolome-drug association atlas, details in Supplementary Table 11), the association of AD and β-hydroxybutyrate is still robust (Supplementary Table 12).

### The associations of AD and energy metabolism-related metabolites in the brain tissue are different as in the periphery

AD is a neurodegenerative disease of which most pathological manifestations are expressed in the brain. Blood-brain barrier may cause the metabolite association in the brain different as the observation in the periphery. Thus, we further explored the association of the identified energy-related metabolites levels (including valine, and β-hydroxybutyrate, acetone not available) in the brain tissue from an aging cohort, the Religious Orders Study and Memory and Aging Project (ROS/MAP), with AD related traits. Interestingly, we identified a different result in the brain as in the periphery: in the brain, the levels of valine are significantly positively associated with all the tested AD related traits, including the levels of tangles, amyloid, cognitive decline, diagnosis of AD and multiple neuropathological variables; while the association of β-hydroxybutyrate levels and AD was not identified in the brain (p>0.05) (Supplementary Table 13).

## Discussion

In this large-scale metabolomic study using 249 NMR-based peripheral metabolites in UK Biobank, we identified 56 metabolites that were significantly associated with incident dementia. We found a strong overlap (82%) in the metabolites significantly associated with the risk of dementia and the metabolites with reaction time in those who did not develop dementia during follow-up. Furthermore, we found that the metabolite signatures of incident dementia significantly correlated with those of total brain volume. AD and VaD were associated with different metabolite signatures, indicating that β-hydroxybutyrate, acetone and valine are associated with AD exclusively. We found that the metabolites related to AD were determined by age and *APOE* genotype and that valine showed a non-linear association across the studied age range, paralleling observations for BMI. Moreover, it is of interest that the associations of AD with valine and β-hydroxybutyrate in the brain tissue are different as their associations in the periphery, which implies the key role of transports on blood-brain barrier in regulating the energy metabolism of AD.

Comparing our findings to those published earlier,^10,12^ we found that the effect estimates of our study and the largest study published^10^ to date are in agreement (Supplementary Figure 5), increasing confidence into these metabolite association. Moreover, the major novel finding of the present study is that we found strong evidence pointing the role of energy metabolism in the development of AD which occurs a decade before its diagnosis based on symptoms: the levels of β-hydroxybutyrate, acetone and valine in blood associate with future AD and also its early pathology (reaction time and total brain volume) in those who did not develop dementia during follow-up. This adds population-based evidence to the previous findings that the disturbance in energy metabolism is an early feature of the AD^18,24,25^ that may be relevant for prevention and treatment.

We are the first study found strong evidence in the population level that in the patients who developed dementia during the follow-up, Acetyl CoA metabolism in the periphery is shifting to alternative energy source when the glucose oxidation and breakdown is less inefficient (Figure 5)^26^. We identified positive association of dementia and AD with ketone bodies (β-hydroxybutyrate and acetone), which indicates a shift from glucose to ketone bodies as the alternative source of energy in case of energy shortage in the brain (Figure 5). The other metabolites identified to be associated with dementia also pointing to energy metabolism: citrate, of which higher levels in the periphery were observed in those who developed dementia, is one of the key metabolites in TCA cycle – the main cellular process to supply energy for the whole body; the low levels of circulating amino acids, including valine and total BCAAs, triglycerides, cholesterol and fatty acids in the future dementia patients also provide evidence as these metabolites can be used as indirect substrates of the TCA cycle through Acetyl CoA metabolism (Figure 5). Our findings are in line with the increasing evidence for mitochondrial dysfunction and the reduction of cerebral metabolic rates for glucose in the patients with AD^21^. Under normal conditions, the brain relies on catabolism of glucose for adenosine triphosphate (ATP) production. Although patients with a high risk of AD have high levels of glucose in the periphery^9^, cerebral metabolic rates for glucose is reduced, particularly in *APOE4* carriers^19^. Astrocytes in the brain are ketogenic and can produce ketone bodies for the neurons as alternative fuel^20^. Alternatively, the liver can produce ketone bodies which can be transported across the blood-brain barrier based on the concentrations in the circulation and be up-regulated by increased energy requirement (liver-brain axis; Figure 5)^20^. Astrocytes expressing *APOE4* are less inefficient at lipid transport^27^ and an early morphological atrophy of astrocytes was observed in the early stage of AD^28,29^, which implies that the capacity of a ketogenic function of astrocytes may be decreased in the *APOE4* carriers and in the people under the early stage of AD. This leads to a higher energy requirement of brain from the periphery and triggers the up-regulation of ketone body transport; thus more ketone bodies from the liver are produced to transport to the brain through circulation. This hypothesis is partially supported by our findings that the association of β-hydroxybutyrate and AD was identified in the periphery but not obvious in the brain. It is also supported by previous findings from the interventional studies which reported that ketogenic diet benefits the cognitive function. This is the first study providing evidence for a molecular cross-talk between the liver and brain in a large-scale population level, in which ketone bodies produced in the liver enter the blood circulation in people who carry *APOE4* variant or are in the early stage of AD. β-hydroxybutyrate constitutes up to 70% of the ketone bodies and the production of β-hydroxybutyrate in the liver is a general compensation mechanism in the body to protect the brain and other organs from damage in case of energy shortage^30,31^. To exclude the possibility that β-hydroxybutyrate is increased in the periphery because of co-morbidities in patients with AD at midlife, we conducted a sensitivity analysis adjusting for known co-morbidities, drugs and biomarkers. None of these explained the association between β-hydroxybutyrate and AD. Moreover, the huge sample size in our study allowed us to examine the association of metabolites in the periphery with age across a large age range and had enough power to shed light on the biological relevance of the change of metabolite levels. This is the first study that explored the levels of ketone bodies in blood over a long age range. We found that the levels of ketone bodies linearly increase over age and that β-hydroxybutyrate is consistently elevated in *APOE4* carriers across the age range (37 to 73 years). This finding is compatible with the view that *APOE4* carriers, who have a higher genetic risk and earlier onset of AD, have a less inefficient function of astrocytes and a relatively increased need for alternative energy sources in the brain already earlier in life compared to non-carriers.

**Figure 5.**
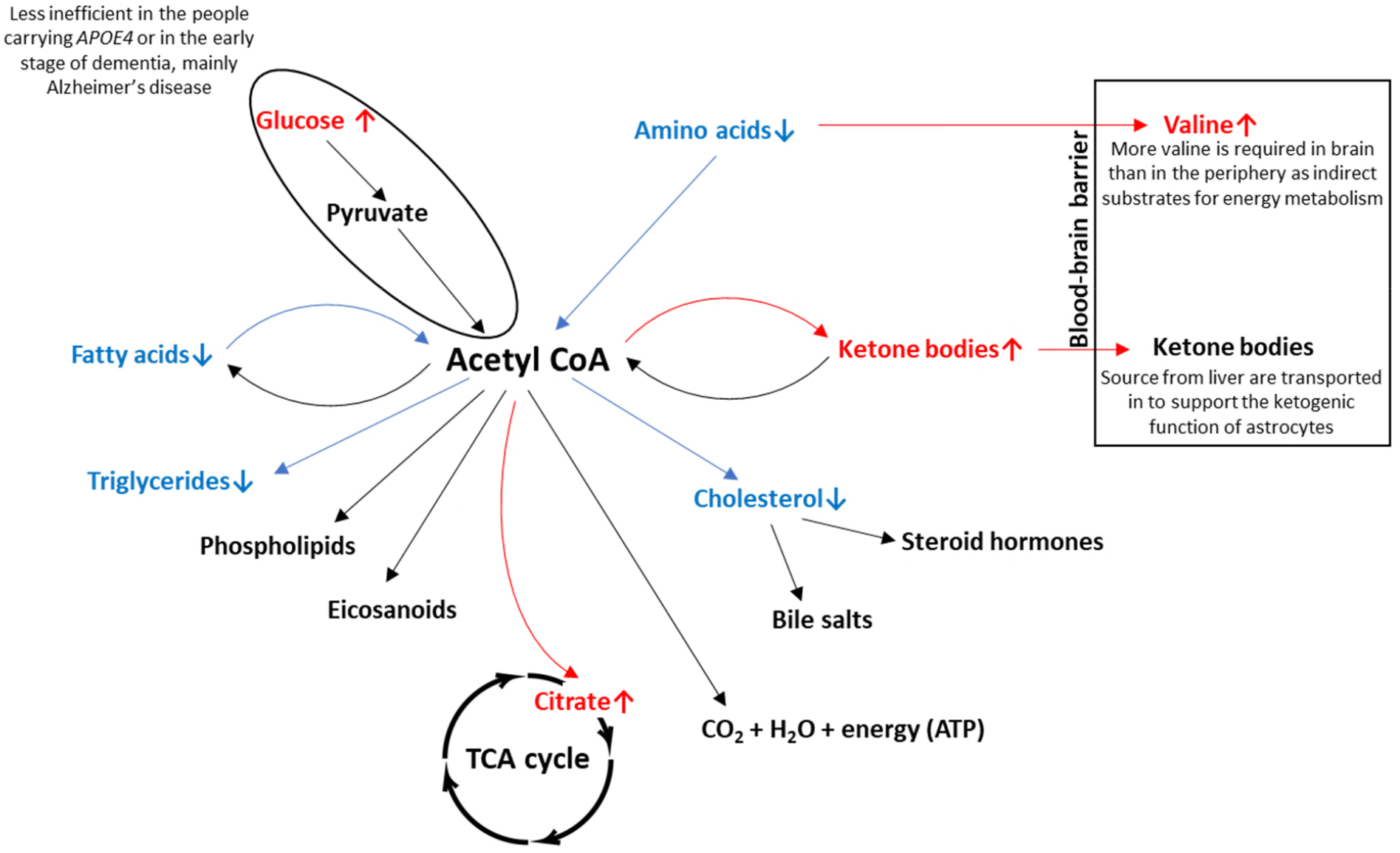
Integration of the metabolite associations for dementia in the regulation of Acetyl CoA and energy metabolism The figure shows that when the energy metabolism through glucose oxidation and breakdown is less inefficient in the *APOE4* carriers or the patients in the early stage of dementia (positive association of glucose and dementia in the periphery), while the levels of amino acids, fatty acids, triglycerides and cholesterol all decrease to support the alternative source of energy (increased citrate and ketone bodies). More valine is required in brain than in the periphery as indirect substrates for energy metabolism, thus the limited valine from food intakes is pumped to brain, leading to even less valine in the periphery. On the other hand, liver produces more ketone bodies and are transported to the brain to support the ketogenic function of astrocytes as alternative energy source. TCA cycle: tricarboxylic acid cycle. ATP: Adenosine triphosphate.↑ metabolite in blood positively associates with dementia; ↓ metabolite in blood negatively associates with dementia. → increased transport or transfer; → decreased transport or transfer.

The other interesting novel finding is the distribution of valine levels in blood across the age range which shows a very different pattern as ketone bodies: the levels of valine were increased with age up to 57-58 years. After age 58, valine levels were decreased in *APOE4* and *APOE33* carriers but not obvious in *APOE2* carriers. The association between low levels of valine in the periphery and an increased risk of AD or cognitive dysfunction is well established and has been reported in many studies^10,32,33,14–16,34^. The negative association of valine and BCAAs in blood and dementia is independent from the effect of cardiometabolic disorders, as diabetes, cardiovascular diseases and obesity are associated with higher levels of valine in blood^13,35–38^. We found that the relation of valine levels in blood with age in *APOE4* carriers is very similar to what we observed for BMI in relation to age. Consequently, BMI and valine in blood were strongly correlated across the whole age range we studied. Combining our findings of the decreased levels of fatty acids and amino acids in the future dementia participants, the decreasing BMI after plateau at age 60 years in *APOE4* carriers may be explained by the fact that if glucose fails as an energy source in the brain and perhaps elsewhere like muscle^17,21^, fatty acids derived from stored adipose tissue are the first alternative energy sources, thus reducing weight and BMI. If fatty acids are no longer available, amino acids derived from muscle tissue may be used for energy production, further reducing the BMI^39^. On the other hand, we identified an increased level of valine in the brain tissue of the participants with a risk of AD, which is in opposite as their association in the periphery out of blood-brain barrier^10,32,33,14–16,34^. This may be explained by the impaired astrocytes in the early stage of AD, or less inefficient astrocytes in the *APOE4* carriers^27^, resulting in a decreased BCAA metabolism in the brain^40^. Thus, besides ketone bodies, more valine in the periphery should be transported to the brain to support the energy metabolism of brain. However, unlike ketone bodies which can be produced by liver, the source of valine in the whole body is limited. Thus, in the people with high risk of AD, an increased valine in the periphery is pumped into brain through blood-brain barrier^41^, resulting in an increased level of valine in the brain and decreased level of valine in the periphery (Figure 5). This also explains the previous finding of randomized clinical trial that BCAA diet improves cognitive function in the people in 60s^42^. Moreover, our results implies that *APOE4* carriers will gain more benefits from the BCAA diet or supplementation, especially valine supplementation, than the others. In summary, our study suggests that while β-hydroxybutyrate in the periphery increases over the complete age range studied in *APOE4* carriers, valine in the periphery only increases up to a certain age (around 57-58 years), after which the levels start to decrease with age, which may due to the impaired astrocytes in the early stage of AD and the less inefficient astrocytes in the *APOE4* carriers. The decrease in valine in blood and the involuntary loss of weight may be biomarkers of the collapse of energy balance more in the carriers with *APOE4* variant and in the people in the early stage of AD than the others^43^.

This study has various limitations. Firstly, the diagnoses of AD and VaD were only available in a limited number of patients and not always precise^44^, but we still provided evidence that metabolite signatures are different between AD and VaD. Secondly, the MRI measurements were not conducted at the same time point as the measurements of the metabolites (on average 9.3 years after the blood collection). Considering that dementia is a neurodegenerative disease with very slow progression, we assume that the MRI phenotypes measured later could still respond to the brain pathological changes at baseline. Thirdly, the Nightingale platform is a cardiometabolic platform which only covers a limited metabolic spectrum. Despite these limitations, the strength of our study is the prospective design in which metabolites were measured before the onset of disease. The in-depth phenotyping facilitates the integration of genetic, epidemiological, clinical and pharmacological exposures.

The large sample size allowed us to study the effect of metabolites across a large age range by *APOE*. Finally, we integrated the findings of dementia and its early pathology by studying the effects of each metabolite and their joint effects as PCs. The use of PCs overcomes the potential false positive associations caused by the high correlation of metabolite measures from abundant metabolite classes (Supplementary Figure 2), e.g., the large number of metabolites in the VLDL class.

In conclusion, we identify a large number of metabolites associated with dementia, through which we emphasize the importance of energy metabolism in the development of AD. We find variant levels of valine and ketone bodies across age range and *APOE* genotypes and different associations of AD and valine in the brain tissue and in the periphery, which provides strong evidence that the onset of AD in *APOE4* carriers is regulated by the impaired energy balance in the brain.

## Methods

### Study design and participants

We performed a prospective, population-based cohort study based on the UK Biobank dataset^45^, which comprised more than 500,000 participants aged from 37 to 73 years during recruitment (2006 to 2010) with non-fasting blood sampling. Among them, a random subset of 118,466 individuals was characterized using the high-throughput ^1^H-NMR metabolomics (Nightingale Health, Helsinki, Finland) platform. The participants were registered with the UK National Health Service and from 22 assessment centres across England, Wales, and Scotland using standardized procedures for data collection which included a wide range of questionnaires, anthropological measurement, clinical biomarkers, genotype data, etc. The imaging data were collected during the third instant follow-up with ∼50K subjects. The participants’ hospital inpatient records and death registration were obtained and updated frequently. The updated data until September 2020 were used to define incident diseases in the current study. After excluding the samples that failed the Nightingale Health quality control metrics (see below), the prevalent dementia cases, and the individuals with missing information on fasting time, we ended up with 118,021 individuals in the current analysis. All participants provided electronically signed informed consent. UK Biobank has approval from the North West Multi-centre Research Ethics Committee, the Patient Information Advisory Group, and the Community Health Index Advisory Group. Further detail on the rationale, study design, survey methods, data collection and ethical approval are available elsewhere^45^. The current study is a part of UK Biobank project 30418 and 54520.

Brain tissue used in this study was obtained from the autopsy collections of ROS/MAP^46^, which is a longitudinal cohort study of aging and dementia in elderly nuns, priests, brothers and lay persons. Brain tissue used in this study was obtained from the autopsy collections under brain donation programs with standardized protocal^46,47^. Tissue was from the dorsolateral prefrontal cortex (Brodmann Area 9 where available) or temporal cortex and precuneus regions where indicated. The Postmortem neuropathological evaluation, extent of spread of neurofibrillary tangle pathology and neuropathologic diagnoses were made in accordance with established criteria and guidelines^25^. As detailed case metadata with age, sex, post-mortem interval (PMI), cognitive function, APOE genotype, neuropathological criteria and disease status was available in previous publication^25^. All procedures and research protocols were approved by the corresponding ethical committees of our collaborator’s institutions as well as the Institutional Review Board (IRB) of Columbia University Medical Center (protocol AAAR4962). More details can also be found in the website of Rush Alzheimer’s Disease Center (RADC; https://www.radc.rush.edu/).

### Definitions of dementia and its related endophenotypes

We defined incident dementia and the major subtypes and onset date based on the previous outcome adjudication guidelines in UK Biobank^48^. In brief, the diseases were based on the self-reported illness (field 20002), or the ICD codes from hospital admission electronic health records in the primary or any secondary causes and/or death register. As the subtypes of dementia were not specified in the self-reported records, they were defined based on hospital admission and death register records only. The earliest recorded code date of diseases was used as the date of disease diagnosis. Prevalent cases were defined as the participants with the disease diagnosis date earlier than the first assessment date, reported in the first time self-reported illness, or using specific indicating anti-dementia drugs for dementia (i.e, donepezil, rivastigmine, galantamine). They were excluded from the survival analysis. The censor date was defined by either the first recorded date of dementia, death date or the end of the digital recording date, whichever happened first. Two baseline variables of cognitive function were used based on previous publication^49^: 1) fluid intelligence score based on the unweighted sum of the number of correct answers given to the 13 fluid intelligence questions (n=36,715), and 2) reaction time based on mean time to correctly identify matches in the cognitive test (n=114,746). The magnetic resonance imaging (MRI) was captured on the median 9.3 years after the collection of the blood samples for the NMR measurements. Three common dementia-related brain MRI regions (average of left and right sides) were selected, of which white matter hyperintensities was from T2 weighted brain MRI (n=9,191), and hippocampus volume and total brain volume were from T1 structural brain MRI (n=9,585). If the variable was measured with both left and right sides, the mean was calculated. To keep consistent, we further log-transformed the MRI data and followed by a standardized scaling as we did for the metabolites.

In ROS/MAP, we studied two direct neuropathological variables, including overall amyloid levels and the levels of tangle density which were measured as the mean of the eight brain regions tested. Three derived neuropathological variables were calculated: global neuropsychiatric scores based on the summary of AD pathology derived from counts of three AD pathologies: neuritic plaques, diffuse plaques, and neurofibrillary tangles; AD diagnosed based on the National Institute on Aging (NIA) Reagan score^50^, and neuropsychiatric diagnosis based on Braak and CERAD scores^51^. Moreover, we also studied three variables reflecting cognitive function: the final consensus cognitive diagnosis at the time of death based on all lifetime clinical data, the global cognitive function based on the last time point of the lifetime cognitive abilities from the results of an average of 19 cognitive tests, and person-specific rate of change in the global cognition variable over time based on lifetime cognitive abilities (change over time). More details can also be found in the website of Rush Alzheimer’s Disease Center (RADC; https://www.radc.rush.edu/).

### Definition of covariates in UK Biobank

The general covariates considered in the analysis included baseline age, sex, BMI, fasting time, assessment center, technical variables during the NMR measurement, i.e., batch and spectrometer, and additional ethnicity, smoking status, alcohol intake frequency, education and drug usage from the verbal interview. BMI was calculated from measured weight and height squared. Fasting time was defined as the interval between the consumption of food or drink and blood sample(s) being taken and log-transformed. Ethnicity was categorized to White, Asian (excluded Chinese), Black, Chinese, mixed and others. Smoking status was categorized as never, previous and current. Alcohol intake frequency was categorized to 1) daily or almost daily, 2) three to four times a week, 3) once or twice a week, 4) less than once a week. Education was categorized to 1) College or University degree, 2) A levels AS levels or equivalent, 3) CSEs or equivalent, 4) NVQ or HND or HNC or equivalent, 5) O levels GCSEs or equivalent, 6) Other professional qualifications, and 7) none of the above based on the highest qualification. If the answer was “prefer not to answer”, missing would be used instead. Medication status was based on the medication codes collected from the verbal interview which were further coded to Anatomical Therapeutic Chemical (ATC) codes^52^. Medications with less than 20 users in the UK Biobank population were ignored during the coding. The medications considered in the covariates were selected based on our previous publication^23^, including five anti-hypertensives (C08, C09, C07, C03 and C02), anti-diabetes (metformin and other anti-diabetes under A10), lipid-lowering drugs (C10), digoxin (C01AA), antithrombotic (B01AC06), proton pump inhibitors (PPI, A02BC), and also 18 drug categories involved in the central nervous system based on 4 digits of the ATC codes. The procedures of other biomarkers used in the current paper are described by UK Biobank^53^. The definitions of other diseases are described in Supplementary Table 14.

### Imputation of missing values in the covariates

Fast imputation of missing values by chained random forests was performed through the R package *missRanger* to impute the missing values for the shared covariates, including smoking status, BMI, alcohol intake frequency, education and ethnicity. The information used in the imputation included baseline age, sex, smoking status, pack-years of smoking, alcohol intake frequency, physical activity from International Physical Activity Questionnaire (IPAQ) groups^54^, ethnicity, BMI, education, blood pressure and waist-hip ratio. In brief, the large matrix was imputed with maximum of ten chaining interactions and 200 trees and weighted by the number of non-missing values; three candidate non-missing values were selected from in the predictive mean matching steps.

### Genotype measurement in UK Biobank

UK Biobank genotyping was conducted by Affymetrix using a bespoke BiLEVEL Axiom array for ∼50K participants and the remaining ∼ 450K on the Affymetrix UK Biobank Axiom array. As the two arrays are broadly comparable with over 95% overlap in assessed gene variants, they were combined. All genetic data were quality controlled and imputed by UK Biobank. The *APOE* gene (alleles *APOE2, APOE3, APOE4*) was directly genotyped and defined by 2 single-nucleotide polymorphisms (SNPs), rs429358 and rs7412. Detailed information on the genotyping process and technical methods are available online^55^. We followed the UK Biobank’s recommendation to exclude the participants who had failed quality control, significant missing data or heterozygosity. The detailed *APOE* genotypes were used as a covariate in the regression models. Participants carrying *APOE24* were excluded when focusing on the effect of *APOE*2 or *APOE4*, considering its combined effect of *APOE2* and *APOE4*. Participants carrying *APOE33* were used as the reference group.

### Metabolites measurement

The metabolites in UK Biobank were measured in plasma using targeted high-throughput ^1^H-NMR metabolomics platform (Nightingale Health Ltd; biomarker quantification version 2020)^22^ which includes 249 metabolite measures simultaneously quantified. They include clinical lipids, lipoprotein subclass profiling with lipid concentrations within 14 subclasses, fatty acid composition, and various low-molecular-weight metabolites such as amino acids, ketone bodies and glycolysis metabolites quantified in molar concentration units. Of them, 165 are directly measured metabolites and 84 are derived measurements. A summary of the metabolites assessed is shown in Supplementary Table 1. The technology is based on a standardized protocol of sample quality control and sample preparation, data storage and automated spectral analyses^56,57^. The values below the detectable threshold were imputed by Nightingale Health and were included in the current analysis. Technical details and epidemiological applications have been reviewed^56,57^. The data obtained from the baseline sampling were used. For the samples with repeated measurement, one of the values was extracted at random. The metabolite values which were suggested to be technical errors in the quality control provided by Nightingale Health during the measurement procedure were treated as missing. Considering the various distributions of metabolites and to make our results comparable with previous findings, we did a natural logarithm transformation of each metabolite after that the zero values were replaced by the lowest value except for zero. Then we scaled these transformed values to standard deviation units.

Metabolomic data used in the current study was generated in 500 samples with brain tissue using Metabolon Precision Metabolomics platform which used an ultrahigh performance liquid chromatography-tandem mass spectrometry (UPLC-MS/MS) system (Metabolon, Inc., Morrisville, USA). The levels of valine and β-hydroxybutyrate were studied. More details can be found in previous publication^58^.

### Statistical analysis

All analyses were performed in R statistical software (version 3.6.2) and the two-tailed test was considered.

#### Metabolite association analysis

In the UK Biobank, we used mixed Cox proportional hazards models with ethnicity as random effect^59^ to estimate the relation between the metabolite levels at baseline and the risk of incident dementia or major subtypes during the follow-up. For analysis conducted in individuals in those with a European background only, a general Cox Proportional Hazards model was used. The proportional hazards assumption of metabolites on dementia was verified for all analysis (Supplementary Table 5 and 8). A false discovery rate (FDR) of 0.05 was used to identify significance. The imbalance in age between those who did and did not develop dementia resulted in a violation of the Proportional Hazard assumption (Supplementary Figure 6). Limiting the analysis to those 60 years or older resolved the problem. For the association analysis, we considered three models with an increasing number of covariates. The basic model adjusted for age, sex, BMI, fasting time, assessment center, and technical variables during the NMR measurement, i.e., batch and spectrometer. The discovery model was further adjusted for smoking status, alcohol intake frequency, education and drugs which were previously found to be associated with the majority of the Nightingale metabolites^23^. Since *APOE* is a genetic determinant of both dementia and metabolomics, we also performed a sensitivity analysis with additionally adjusted for *APOE* genotypes.

Linear mixed regression models were used to estimate the association of metabolites (or PCs) with *APOE*, where metabolites were used as dependent variables (adjustment according to the discovery model). We further associated the metabolites with endophenotypes, modelling the endophenotypes as independent variables (adjustment according to the discovery model). As metabolite spectra may change as a consequence of disease, these analyses were performed in the participants who did not develop dementia in the follow-up. Since the imaging data were collected almost ten years after the baseline enrolment, the associations of metabolites and MRI traits were measured in the participants aged 50 or older at baseline. Following the protocol from UK Biobank^60^, the head size and all the head position coordinates were also adjusted when analysing brain imaging data. For generating heatmaps, we used hierarchical clustering with the default setting in *corrplot()* function in R.

The association analysis of valine and β-hydroxybutyrate was studied by linear regression with adjustment for gender, BMI, post-mortem interval, age at death, education and *APOE* genotypes. All the AD-related traits are derived as positively associated with the risk of AD before doing the association with metabolites.

#### Integrating metabolite signatures of dementia and endophenotypes

Since the metabolites of the Nightingales platform were highly correlated, we conducted a principal component analysis to create a series of PCs that were independent from each other. We used the first 27 PCs, which explained over 95% of the metabolites (n=249). We tested the correlation of the effect estimates per SE of the PCs between traits using linear regression and Pearson’s correlation.

#### The role of age and APOE on metabolite levels

To determine the impact of age on metabolites across *APOE* genotypes (i.e, *APOE2* carriers, *APOE4* carriers and *APOE33*), we created age-trend plots of the distribution of the levels of metabolites across the full age range (37 to 73 years) in participants who did not develop dementia in the follow-up. We used generalized additive models through the geom_smooth() function in the *ggplot2* package to smooth the lines in the plots. The residuals of metabolites were calculated using the linear mixed regression with ethnicity as the random effect and adjusting for the covariates in the discovery model excluding age. To fit the plots, we performed a regression model with the metabolite levels or its residuals as the dependent variable and the baseline age, the square of baseline age, *APOE* and the interaction of age and *APOE* as independent variables. In the analysis, we replaced baseline age with mean-centred baseline age to control the effect of multicollinearity between age and *APOE*^61^.

## Data Availability

UK Biobank data are available through a procedure described at http://www.ukbiobank.ac.uk/using-the-resource/. Data from ROSMAP are available through
the Rush AD Center Resource Sharing Hub (https://www.radc.rush.edu). All the summary statistics are available in the supplementary tables.

http://www.ukbiobank.ac.uk/using-the-resource/

https://www.radc.rush.edu

## Data Availability Statement

UK Biobank data are available through a procedure described at http://www.ukbiobank.ac.uk/using-the-resource/. Data from ROSMAP are available through the Rush AD Center Resource Sharing Hub (https://www.radc.rush.edu). All the summary statistics are available in the supplementary tables.

## Acknowledgment

We thank contributors who collected samples used in this study and patients and their families, whose help and participation made this work possible. This research has been conducted using the UK Biobank Resource under Application Number 30418 and 54520. The Nightingale datasets have been generated at Nightingale Health.

