## Supplementary Figure for "Longitudinal analysis of UK Biobank participants suggests age and APOE-dependent alterations of energy metabolism in development of dementia"

Supplementary Figure 1 Metabolite associations of incident dementia in multi-ethnic population and European population

A

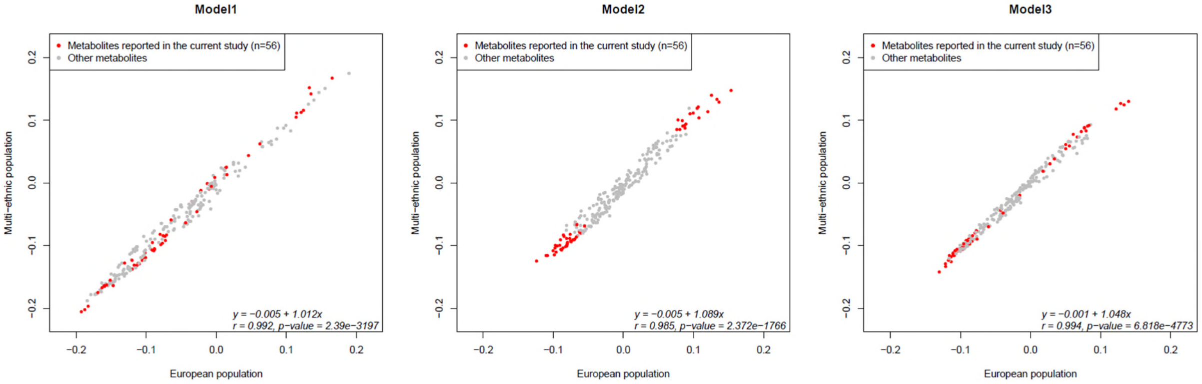


The (Pearson's) correlation of effect estimates of the global metabolite associations between multi-ethnic population and European population. Model1 with adjustment of ethnicity, age, sex, body mass index, fasting time, assessment center, and technical variables during the NMR measurement, i.e., batch and spectrometer**.** **Model2** with adjustment of smoking status, alcohol intake frequency, education and drugs in addition. **Model 3** adjusted for *APOE* in addition.

Supplementary Figure 2 Clustering of the metabolites associated with dementia (n = 56) in the non-demented population.

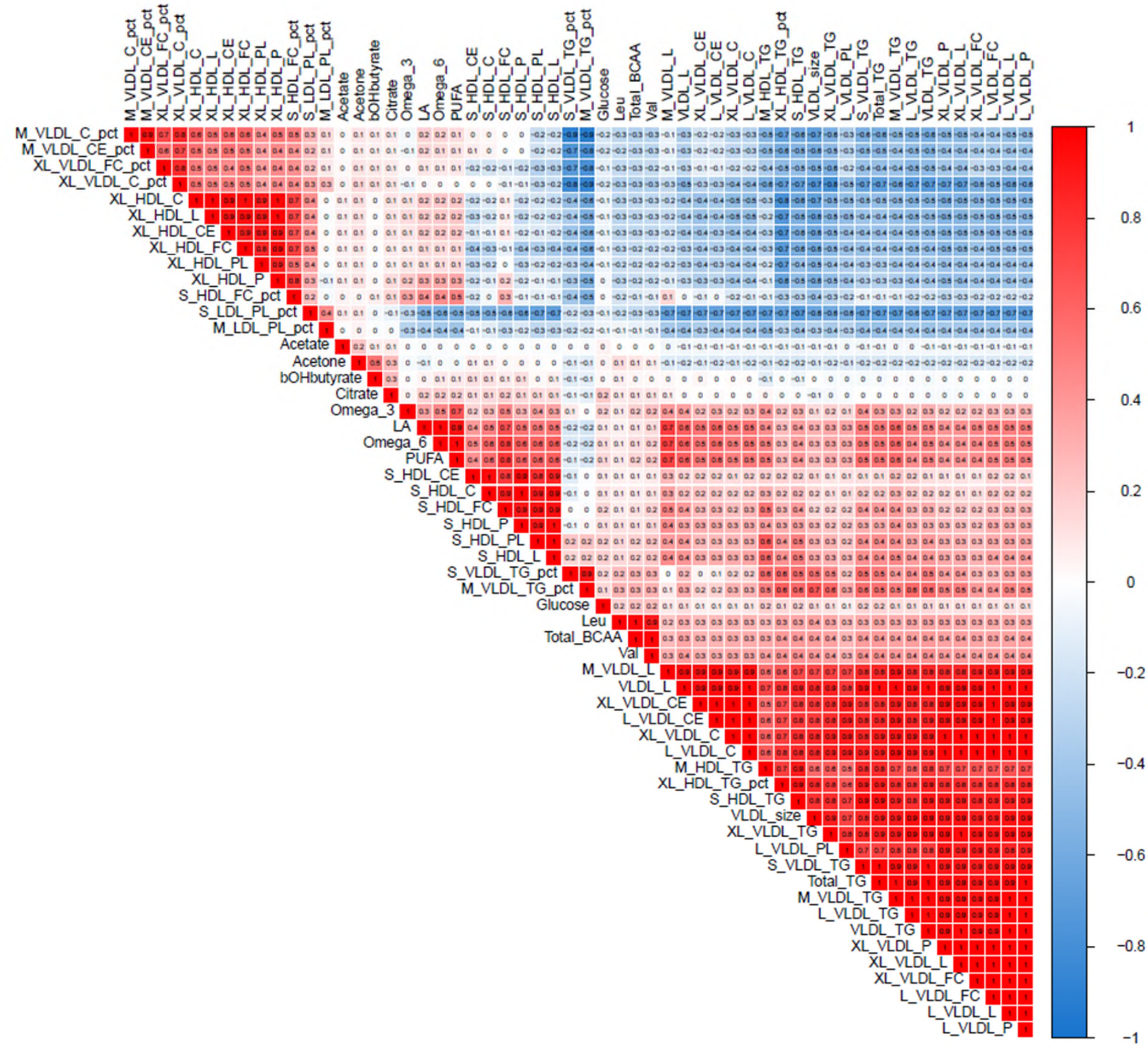


The hierarchical clustering approach was used to determine the orders in the axes. Correlation coefficients are presented in the boxes. The coding of the metabolites are presented in Supplementary Table 1.

Supplementary Figure 3 Sensitivity analysis of integrating metabolite signatures of dementia and white matter hyperintensities and fluid intelligence score in the participants who did not develop dementia during follow-up.

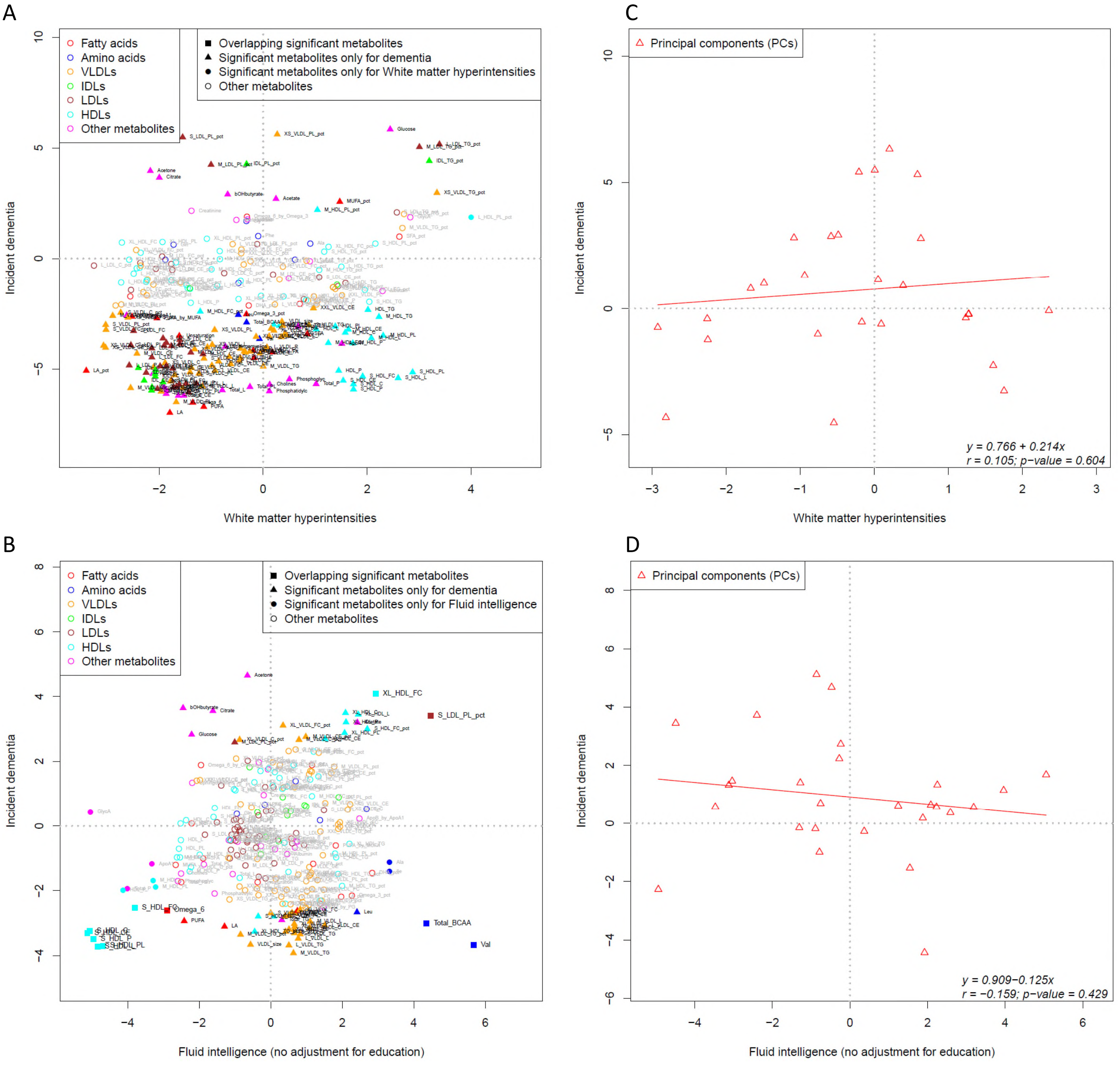


The potential mediating factors of matter hyperintensities and fluid intelligence score were excluded in the metabolites association analyses. For the analysis of white matter hyperintensities, the covariates include ethnicity, age, sex, body mass index, fasting time, assessment center, and technical variables during the NMR measurement, i.e., batch and spectrometer, which excluded the potential cardiometabolic factors included in the discovery model. For the analysis of fluid intelligence scores, the education variable was excluded from the discovery model. The first 27 principal components (PCs) which explain over 95% of the 249 metabolites were used to overcome the high correlation among the metabolites. The effect estimates per standard error with incident dementia and the endophenoypte were presented in the x-axis and y-axis respectively. Linear regression and Pearson’s correlation test were used to fit the linear model of the metabolite association pattern through PCs. bOHbutyrate: β-hydroxybutyrate; Val: valine. The coding of other metabolites are presented in Supplementary Table 1.

Supplementary Figure 4 The distribution of metabolites and BMI over the life course by *APOE* in the population who did not develop dementia, adjusted for covariates in the discovery model.

A B C D

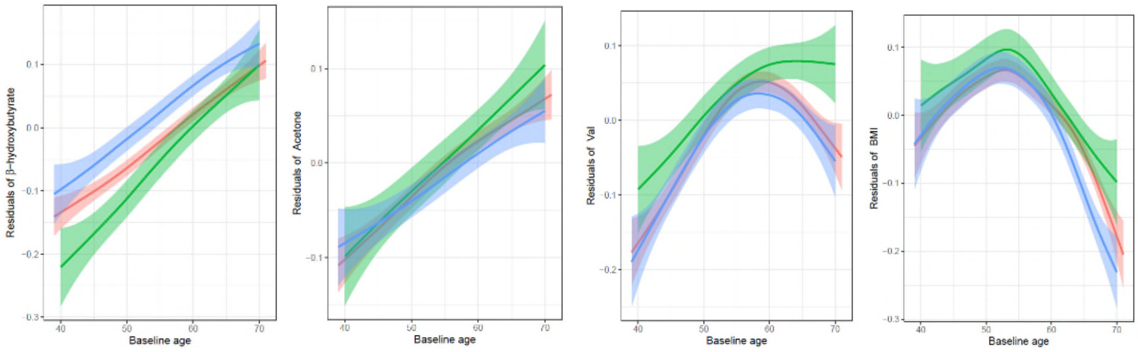


Participants carrying *APOE24* were excluded; participants carrying *APOE33* were used as the reference group (red colour). *APOE2* carriers are presented in green colour and *APOE4* carriers in blue colour. The residuals of metabolites or BMI were calculated by the linear mixed regression with the discovery model (excluded age for metabolites, and excluded age and BMI for BMI) to control the confounding effect of the covariates. Generalized additive model was used to smooth the lines in the figure. Linear regression was performed to model the lines, and showed below. Age: baseline age; age$^2$: the square of baseline age; * interaction. The metabolites are coded based on Supplementary Table 1.

(A) Residuals of β - hydroxybutyrate = - 0.009 + 0.008 × age (p = $1.2 \times 10^{-60}$) - 0.032 × *APOE2* (p = $5.3 \times 10^{-4}$) + 0.044 × *APOE4* (p = $4.8 \times 10^{-10}$) - $5.0 \times 10^{-6}$ × age$^2$ (p = 0.92) + 0.003 × age**APOE2* (p = 0.0064) + 0.0001 × age**APOE4* (p = 0.90)

(B) Residuals of acetone = 0.005 + 0.006 × age (p = $1.0 \times 10^{-30}$) + 0.013 × *APOE2* (p = 0.16) - 0.007 × *APOE4* (p = 0.29) - $5.5 \times 10^{-5}$ × age$^2$ (p = 0.25) + 0.001 × age**APOE2* (p = 0.28) - 0.0007 × age**APOE4* (p = 0.39)

(C) Residuals of valine = 0.039 + 0.004 × age (p = $5.4 \times 10^{-13}$) + 0.041 × *APOE2* (p = $7.2 \times 10^{-6}$) - 0.011 × *APOE4* (p = 0.13) - 0.0006 × age$^2$ (p = $2.3 \times 10^{-40}$) + 0.001 × age**APOE2* (p = 0.34) - 0.0006 × age**APOE4* (p = 0.46)

(D) Residuals of BMI = 0.052 - 0.006 × age (p = $1.0 \times 10^{-39}$) + 0.027 × *APOE2* (p = 0.003) - 0.017 × *APOE4* (p = 0.013) - 0.0008 × age$^2$ (p = $1.3 \times 10^{-59}$) - 0.0001 × age**APOE2* (p = 0.92) - 0.003 × age**APOE4* (p = $8.3 \times 10^{-4}$)

Supplementary Figure 5 Metabolite associations of incident dementia in UK Biobank and in the previous study by Tynkkynen et al.

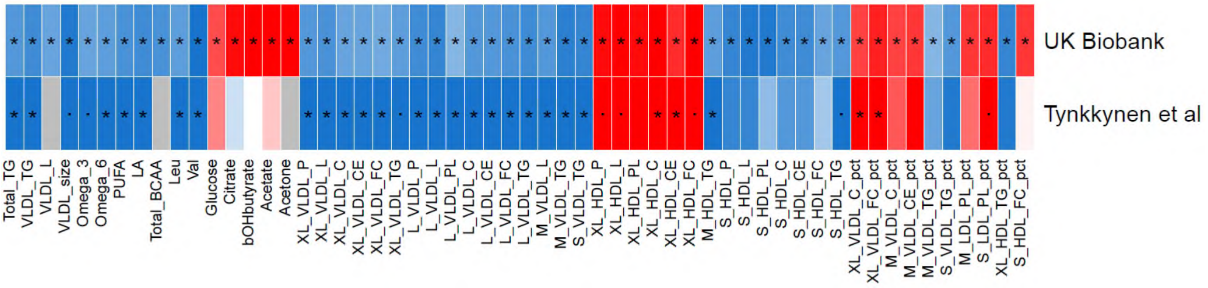


It shows only the significant metabolites (FDR < 0.05, n = 56) identified in the discovery model with adjustment of ethnicity, age, sex, body mass index, fasting time, assessment center, and technical variables during the NMR measurement, i.e., batch and spectrometer, smoking status, alcohol intake frequency, education and drugs. False discovery rate(FDR) adjusted p was calculated in the available metabolites based on the results from Tynkkynen et al's study (n=53). Dot (.) denotes p < 0.05 and star (*) denotes FDR < 0.05. Grey: not available in Tynkkynen et al's study; Red: positive association; blue: negative association; the depth of the color presents the strength of effect estimate per standard error. The coding of the metabolites are presented in Supplementary Table 1.

Supplementary Figure 6 The non - linearity check of the effect of baseline age on incident dementia.

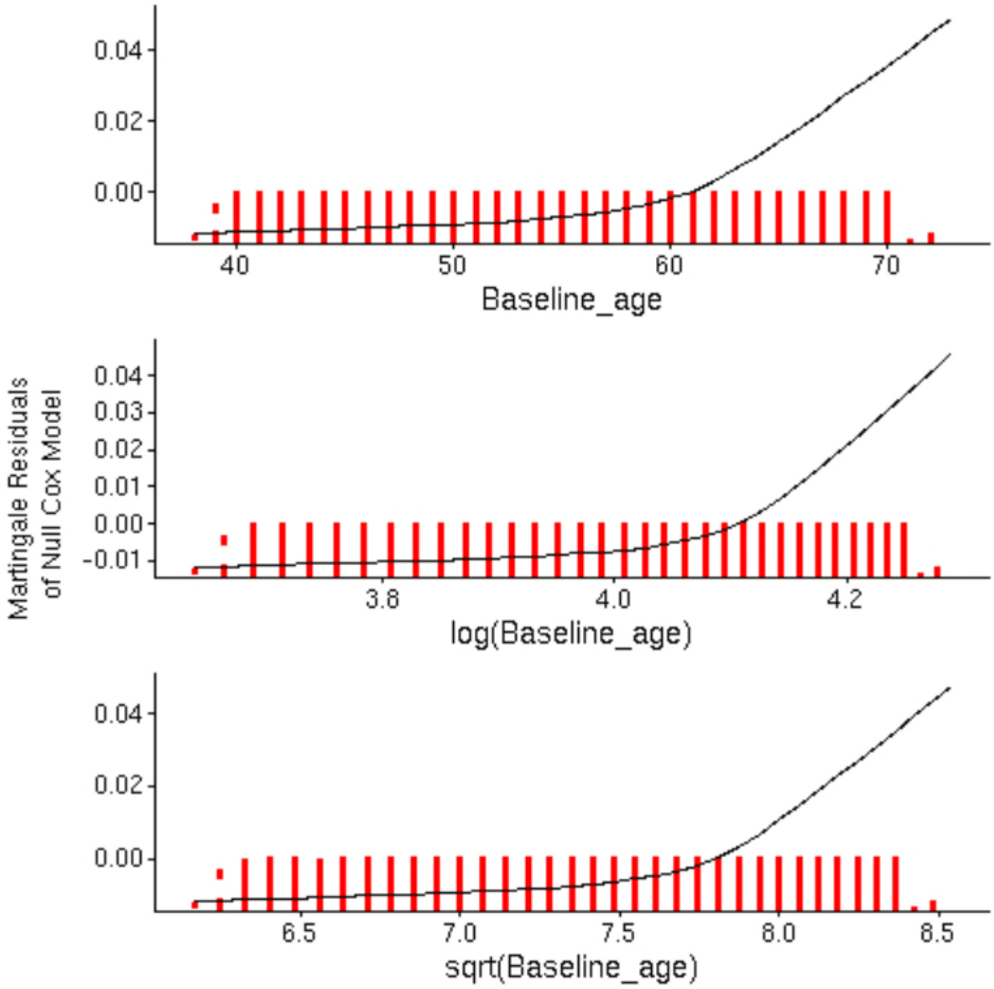


The figure shows the association of baseline age, log - transformed age value and square root - transformed age value on incident dementia under the martingale residuals of null cox model.
